# A synthetic data generation pipeline to reproducibly mirror high-resolution multi-variable peptidomics and real-patient clinical data

**DOI:** 10.1101/2024.10.30.24316342

**Authors:** Mayra Alejandra Jaimes Campos, Stipe Kabić, Agnieszka Latosinska, Ena Anicic, Justyna Siwy, Vinko Dragušica, Harald Rupprecht, Lorenzo Catanese, Felix Keller, Paul Perco, Enrique Gomez- Gomez, Joachim Beige, Antonia Vlahou, Harald Mischak, Davorin Vukelić, Tomislav Križan, Maria Frantzi

## Abstract

Generating high quality, real-world clinical and molecular datasets is challenging, costly and time intensive. Consequently, such data should be shared with the scientific community, which however carries the risk of privacy breaches. The latter limitation hinders the scientific community’s ability to freely share and access high resolution and high quality data, which are essential especially in the context of personalised medicine. In this study, we present an algorithm based on Gaussian copulas to generate synthetic data that retain associations within high dimensional (peptidomics) datasets. For this purpose, 3,881 datasets from 10 cohorts were employed, containing clinical, demographic, molecular (> 21,500 peptide) variables, and outcome data for individuals with a kidney or a heart failure event. High dimensional copulas were developed to portray the distribution matrix between the clinical and peptidomics data in the dataset, and based on these distributions, a data matrix of 2,000 synthetic patients was developed. Synthetic data maintained the capacity to reproducibly correlate the peptidomics data with the clinical variables. Consequently, correlation of the rho-values of individual peptides with eGFR between the synthetic and the real-patient datasets was highly similar, both at the single peptide level (rho = 0.885, p < 2.2e-308) and after classification with machine learning models (rho_synthetic_ = -0.394, p = 5.21e-127; rho_real_ = -0.396, p = 4.64e-67). External validation was performed, using independent multi-centric datasets (n = 2,964) of individuals with chronic kidney disease (CKD, defined as eGFR < 60 mL/min/1.73m²) or those with normal kidney function (eGFR > 90 mL/min/1.73m²). Similarly, the association of the rho-values of single peptides with eGFR between the synthetic and the external validation datasets was significantly reproduced (rho = 0.569, p = 1.8e-218). Subsequent development of classifiers by using the synthetic data matrices, resulted in highly predictive values in external real-patient datasets (AUC values of 0.803 and 0.867 for HF and CKD, respectively), demonstrating robustness of the developed method in the generation of synthetic patient data. The proposed pipeline represents a solution for high-dimensional sharing while maintaining patient confidentiality.

## Introduction

The rapid expansion of omics-based platforms^1^ in combination with advanced imaging, as well as digital health technologies have resulted in the generation of massive amounts of data^2^. Ninety percent of the world’s data has been created within the last two years^3^. These data have the potential to enhance healthcare outcomes^4^ by evolving personalised medicine, especially when analysed via advanced computational analytics and/ or artificial intelligence^5^. To reach this goal, open data sharing and public availability is a pre-requisite, not only as part of the Findable, Accessible, Interoperable, Reusable (FAIR) principles that have been developed^6^, but also as part of good scientific and open science practices to enable reproducibility of the research findings. In reality, the sensitive nature of patient information regularly restricts access and creates substantial challenges for research and development especially within international and/or multi-centric collaborations^7^.

Synthetic data, which are artificially created to replicate real-patient (or real-world) data, offer a promising solution for protecting patient identities. This approach enables the use of a plethora of clinical and molecular data, while adhering to privacy regulations such as the General Data Protection Regulation (GDPR)^8^ in Europe and the Health Insurance Portability and Accountability Act (HIPAA)^9^ in the United States. Several methods for synthetic data generation have been developed, including statistical, probabilistic, machine learning, and/or deep learning approaches^10^.

The applications of synthetic data generation in healthcare has mainly focused on de-identification of clinical and demographical patient data, or re-capitulation of images, and to a lesser extent on simulation, or expansion of molecular data (omics)^10^. For the latter, synthetic data generation algorithms mostly concentrated on either reproducing biological clusters, causal networks and pathways^11–13^, or validating algorithms at the level of visualisation and dimensionality reduction^14^. Moreover, although synthetic data generation algorithms are reported for imputing missing values for large omics datasets (particularly proteomics)^15^, or to adjust for technical noise or intrinsic variability^16^, no synthetic data generation pipeline is in place for mirroring real-patient data that retain association with clinical, demographic and outcome information at high dimensional and high resolution molecular (omics) data level.

Herein, we present a novel approach to generate synthetic data that mirror clinical and high dimensional molecular (peptidomics) datasets (n = 3,881 datasets)^17^, as derived by capillary electrophoresis coupled to mass spectrometry (CE-MS)^18^. For that purpose, we propose a robust statistical/ probabilistic model based on Gaussian copulas^19^.

Copulas are a powerful statistical tool that allows for modelling of the distributions of each variable in a given dataset, as well as the relations between all pairs of variables, by creating a comprehensive modelling of the data. The key technique involves transforming the marginal distribution of each variable into a uniform distribution through application of its cumulative distribution function (CDF)^19^. The Gaussian copula then models the dependencies by capturing the correlation structure of these uniformly distributed variables within a multivariate normal distribution.

In this study, we have utilised copulas and inverse transformation methods to generate synthetic datasets that replicate the associations, dependencies, and marginal distributions observed in the high resolution real-patient data. Transformation methods seem advantageous for computing Bayesian inferences for highly parametrised models and large datasets like peptidomics. When following this approach, the synthetic data are not offsprings of the distinct real-patient data, but rather the approximation of a targeted distribution using multivariate Gaussian methods that is selected to balance accuracy with computational feasibility^20^.

## Results

### Synthetic data generation pipeline to mirror real-patient omics and clinical data

A total of 3,881 real-patient datasets (referred to as ‘training’ set) were extracted from the human urinary peptidomics database^17, 18^. A summary of the patients’ clinical and follow-up data is provided in **Table 1**, including individuals developing a kidney event [MAKE; based on > 40% loss in the estimated Glomerular Filtration Rate (eGFR) or kidney failure; n = 113], those developing a heart failure event (HF; n = 242), or those without developing any of the above during the monitoring follow-up time (NE; n = 3,296; median follow up time was 3.1 years). The real-patient (training) data along with their accompanying molecular (CE-MS) datasets were portrayed into 2,000 synthetic data, based on Gaussian copulas, as described in the Methods section (**Fig. 1**). The specific method was selected as it is not patient-centric, thereby one - to - one patient correlations are completely eliminated. Given the characteristics of the peptidomics matrix^21^, marginal distributions of each feature were estimated following a comprehensive evaluation of different distribution types and their associated parameters. The analysis resulted in lognormal, normal and t-distribution as the optimal distribution types, with the best - fit distribution method that was determined based on the summary of squared errors. To further ensure stability of the algorithm in generating synthetic datasets, and to weigh variability caused by, among others, statistical and functional approximation errors, a bootstrapping approach including 500 iterations was adopted using random sampling^22^. Following a signal threshold for peptides with at least 40% non-zero data in either the individuals with a future event (MAKE or HF), or those without a future event (NE), 2,168 peptides were considered for further statistics. Similarity of data distributions was increased after the bootstrapping approach, resulting in 303 peptides (13.9%) having infinite values for the Kullback–Leibler (KL) divergence^23^, compared to 369 (17.0%) before the bootstrapping approach (**Supp. Fig. SF2**). The distributions of the clinical variables and their correlations were similar within real- and synthetic datasets (**Supp. Fig. SF3**).

**Fig. 1:**
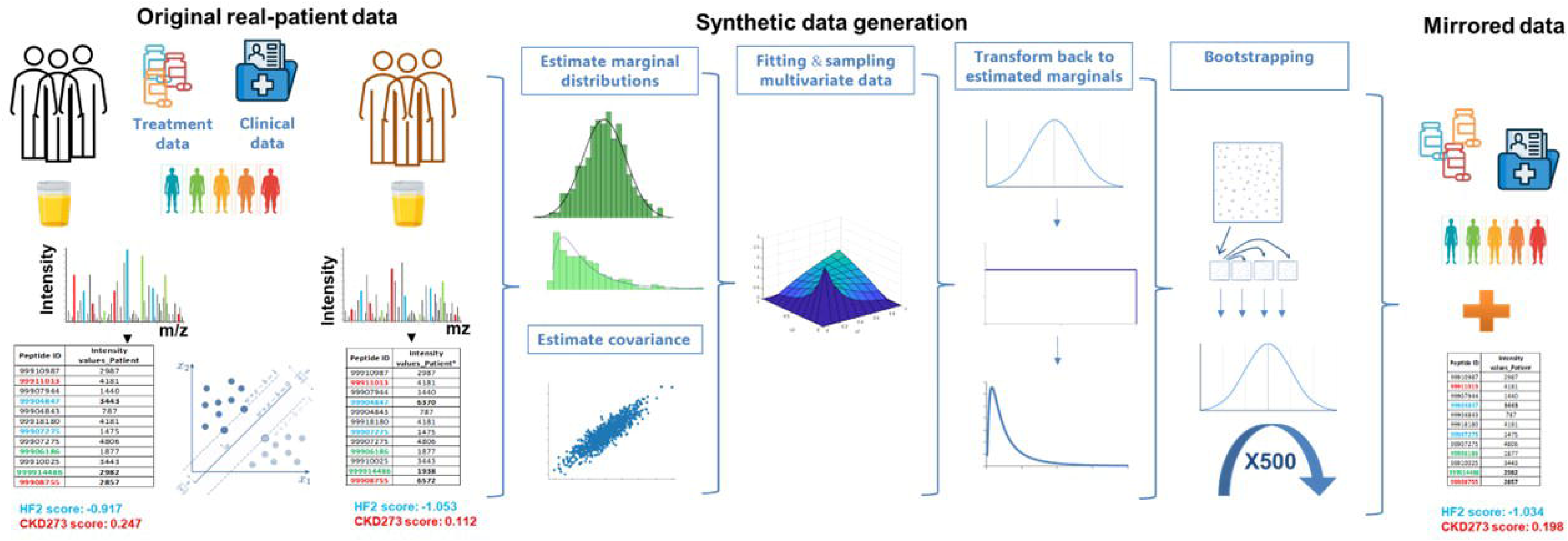
The synthetic data generation pipeline produces datasets highly similar to high resolution peptidomics data from real-patient datasets including clinical, demographic and outcome data.

**Table 1.** Baseline characteristics of study participants and respective variable summary statistics for synthetic and real-patient data.

| Characteristic | N = 3,881 real training patient data | N = 2,000 synthetic mirrored data | P- value |
| --- | --- | --- | --- |
| **Clinic characteristics** |  |  |  |
| Age | 61.0 [54.0, 68.0] | 61.1 [53.3, 67.6] | 0.36 |
| Male Gender | 2338 (60.2%) | 1228 (61.4%) | 0.41 |
| sBP (mm Hg) | 133.0 [123.0, 144.0] | 133.0 [123.5, 144.6] | 0.15 |
| dBP (mm Hg) | 78.5 [72.0, 84.0] | 78.2 [71.9, 84.6] | 0.26 |
| Hypertension | 1575 (40.6%) | 820 (41.0%) | 0.78 |
| Diabetes | 3089 (79.6%) | 1578 (78.9%) | 0.56 |
| eGFR (mL/min/1.73m<sup>2</sup>) | 85.6 [64.2, 97.5] | 85.4 [66.3, 97.9] | 0.42 |
| Time to HF event (years) | 2.2 [0.9, 4.0] | 1.9 [0.8, 3.4] | 0.19 |
| Time to MAKE (years) | 1.2 [1.0, 3.1] | 1.6 [1.0, 2.6] | 0.67 |
| Duration of follow-up non-event (years) | 3.1 [2.2, 4.4] | 3.2 [2.0, 4.2] | 0.29 |
| **Events** |  |  |  |
| HF event | 472 (12.16%) | 242 (12.5%) | 0.98 |
| MAKE | 113 (2.91%) | 58 (3.3%) | 1 |
| Non-event (NE) | 3296 (84.93%) | 1700 (85.21%) | 0.97 |
| Median [IQR]; n (%) |  |  |  |
| Abbreviations: eGFR, estimated glomerular filtration rate (mL/min per 1.73 m<sup>2</sup>); MI, body mass index; sBP, systolic blood pressure; dBP, diastolic blood pressure |  |  |  |

### Multi- level statistical evaluation of the synthetic data generation pipeline

To further assess the validity of the synthetic data generation pipeline, a multi-level verification process was followed based on extended statistical analysis and validation **(Fig. 2**). For this purpose, side - by - side comparisons were performed within the synthetic and the real-patient data, not only through investigating the correlations of single peptides with clinical variables and endpoints **(Fig. 2**), but also at the level of the classification scores when considering well established machine learning models based on support vector machines (SVM; **Fig. 2**)^24–27^. Both were tested in the context of two disease outcomes (MAKE or HF) (**Fig. 2**). At first, summary statistics at the cohort level, demonstrated that the median values of all available clinical variables including demographics, but also blood pressure, eGFR and comorbidities did not differ significantly (**Table 1)**. Even more, outcome data related to the time to develop an HF event or MAKE as well as the follow-up duration were also well reflected in the synthetic datasets (**Table 1**). Besides mirroring the clinical characteristics, the synthetic data also well reflected the peptidomics features, as depicted in dimensionality reduction approaches including principal component analysis (PCA) and Uniform Manifold Approximation and Projection (UMAP; **Fig. 3a-c**). Subsequently the association of peptide abundance with eGFR, a clinical measure of kidney function and a continuous clinical variable available for the entirety of patient datasets, was investigated separately in the synthetic and the real-patient data. Spearman’s rank correlation of each single peptide’s abundance with eGFR in the synthetic and the real-patient data was performed, and the respective rho values were subsequently plotted, revealing high similarity between the synthetic and the real data at the single peptide level (Spearman rho = 0.885; p < 2.2e-308; **Fig.3e; Supp. Table S1**).

**Fig. 2:**
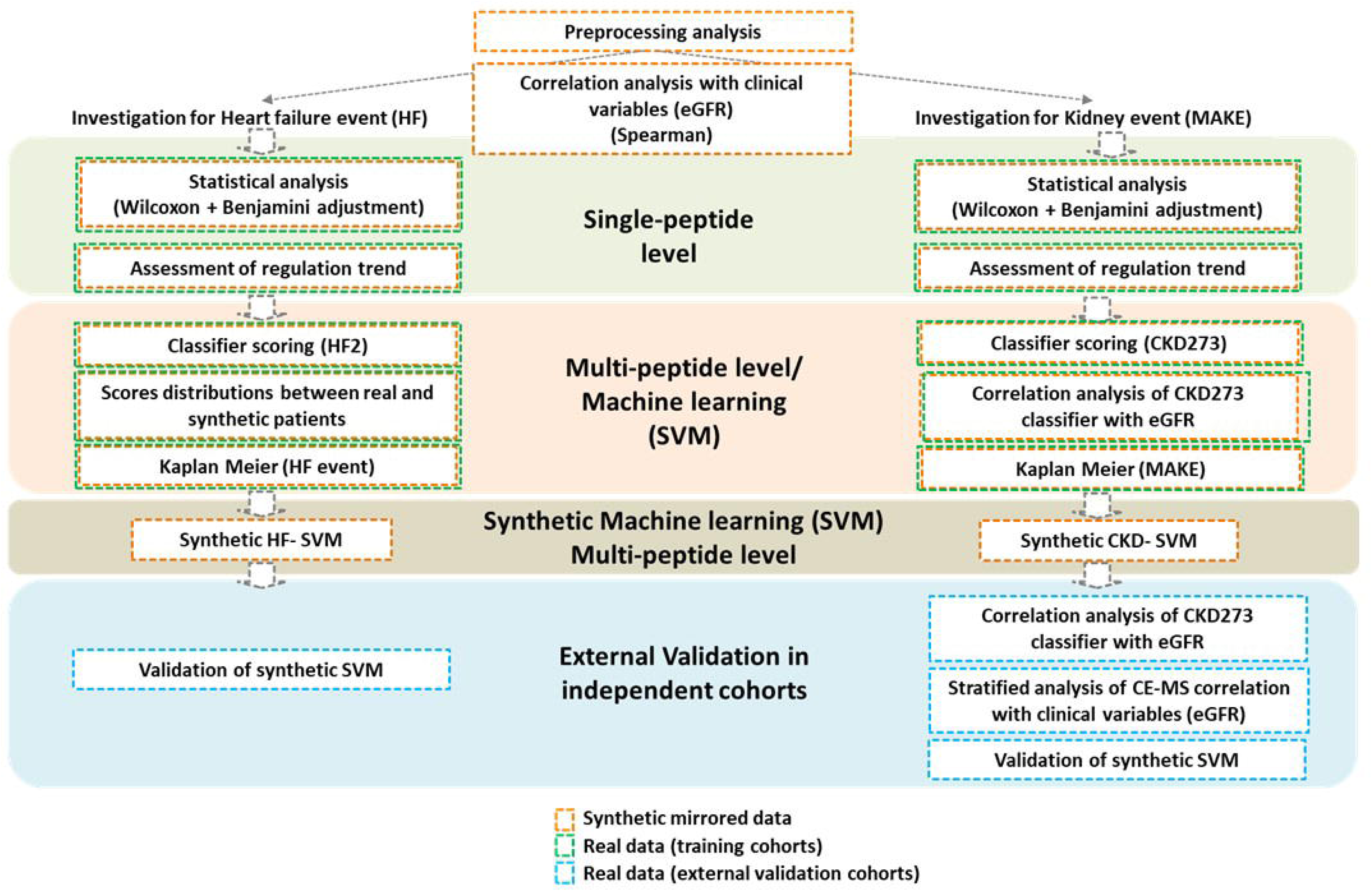
Schematic workflow of the multi-level synthetic data validation including a data matrix of 21,559 peptide derived after CE-MS peptidomics profiling within a real-patient cohort of 3,881 patients. Independent validation was performed in external patient cohorts.

**Fig. 3:**
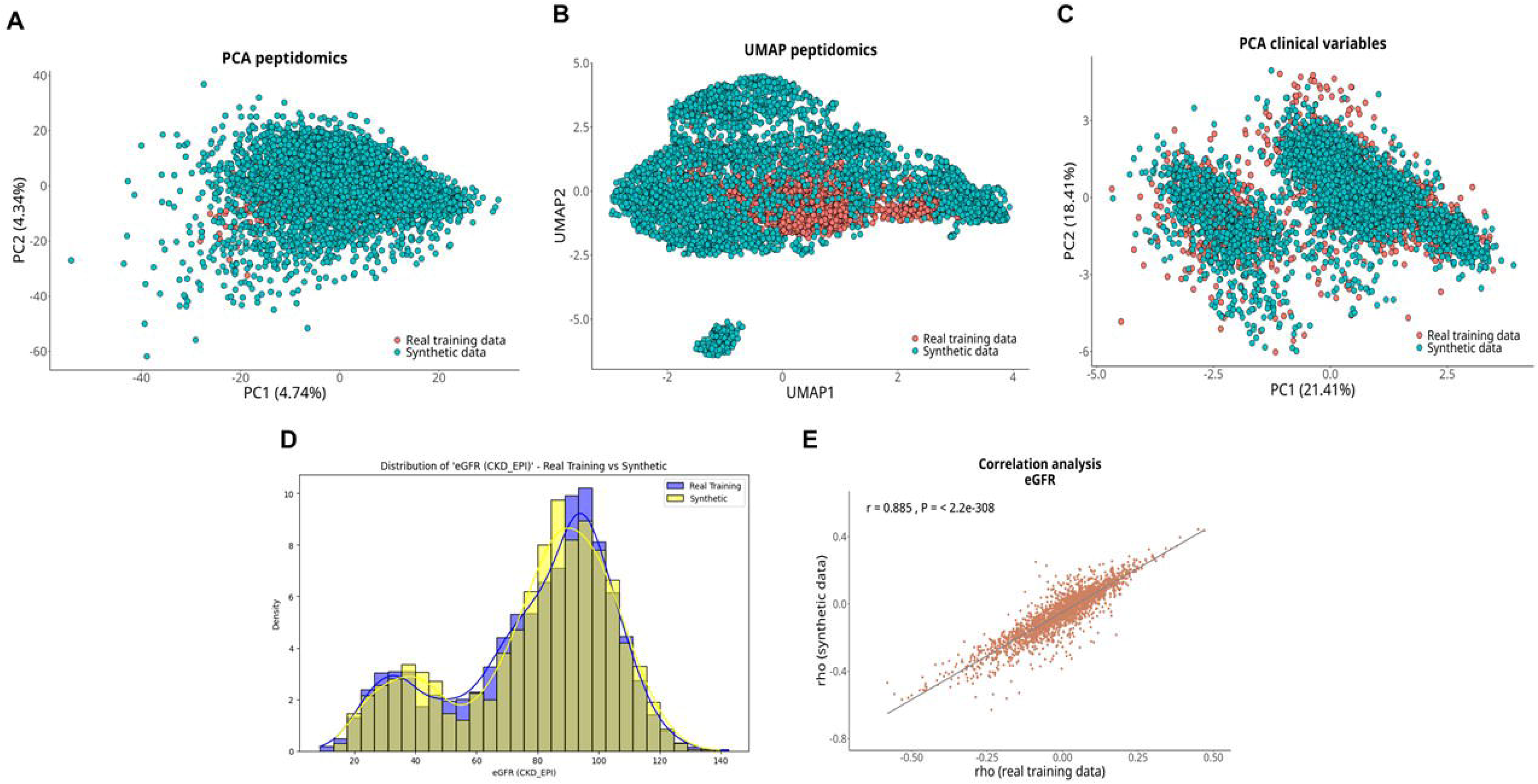
**A-C)** Dimensionality reduction techniques applied in the synthetic and the real-patient (training) datasets demonstrate comparability of the two datasets. **D)** Distributions of the estimates for a highly significant risk factor (continuous variable), like eGFR and **E)** Correlation plots of spearman rho values compared between the real and synthetic data for eGFR. The results confirm the similarity of the generated synthetic data.

### Validation of the predictive associations with heart failure outcome

Considering a future HF event, a proportionally similar number of events in the synthetic and real training data was evident [n_synthetic_ = 242 (12.5%), n_real-training_ = 472 (12.2%) HF events, p = 0.98); **Suppl. Table S2**]. For further comparisons, the synthetic datasets representing individuals that did not develop any event were considered as control group [NE; n_synthetic_= 1,700 (85.2%), n_real-training_= 3,296 (84.9%), p = 0.97)]. A case-control statistical analyses was performed for the peptidomics data from synthetic patients experiencing a future HF event compared to NE group. 1,669 peptides were significantly different in abundance between the two groups in both real and synthetic datasets (**Fig. 4a; Suppl. Table S2**), with consistent regulation trend for the vast majority of them (97.8%) (**Fig. 4b; Suppl. Table S2**). The previously established classifier predicting the HF (HF2 classifier)^28^ was employed to score the real-patient and synthetic datasets. The distribution of the respective scores for the synthetic and the real-patient data supported a very good overlap [(mean of scores for HF_real_: 0.247, mean of HF_synthetic_: 0.270; p = 0.2982) and (mean of NE_real_: -0.334; mean of NE_synthetic_: -0.442; p = 1.0; **Fig. 4c**)], recapitulating the intrinsic discriminatory capability of the classifier in the synthetic dataset. Moreover, the HF2 scores separated in quintiles were predictive of an HF event within both the real patient (hazard ratio (HR) = 2.832 ± 0.047; 95% CI; p = 6.42e-105; **Fig. 4d; Suppl. Table S2**) and the synthetic datasets (HR = 2.809 ± 0.054; 95% CI; p = 1.38e-82; **Fig. 4f; Suppl. Table S2**). The observation was consistently present for both the real-patient and the synthetic datasets (**Fig 4e-f).**

**Fig.4:**
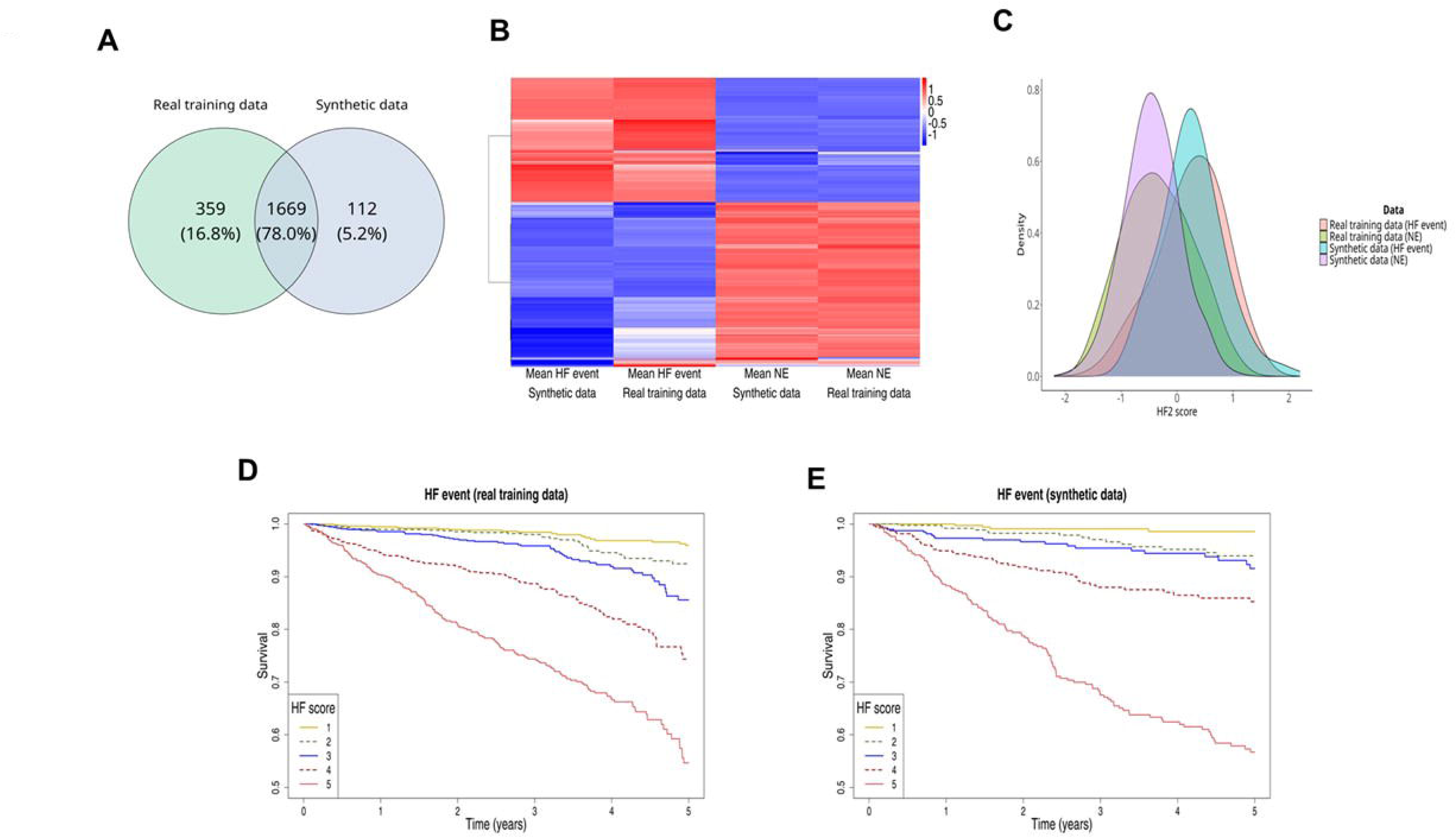
Validation of the synthetic data in heart failure. **A)** Case control comparison between HF and NE patients. **B**) Heat map plot of the most significantly different peptides separating the HF and NE groups, in the synthetic and real-patient data. **C**) Comparative distribution plots of the scores acquired after applying the well-established HF2 classifier in both the synthetic and real data. Kaplan-Meier curves for predicting heart failure event; classifier scores from lowest (1) to highest (5) quintile, for risk of heart failure events as assessed by HF2 in **D**) the real-patient and **E**) the synthetic datasets.

### Synthetic data predicts kidney disease outcome

Clinical comorbidity outcome data on kidney disease (MAKE) were available in the real-patient datasets. The number of MAKE in the synthetic patient datasets was 58 (3.3%), proportionally similar to the real-patient data (n = 113; 2.9%; p = 0.10; **Suppl. Table S3).** Similar to the HF outcome, individuals that did not develop any event were considered as control group [NE; n_synthetic_ = 1,700 (85.2%), n_real-training_ = 3,296 (84.9%), p = 0.97)]. Case-control statistical analyses revealed 745 peptides with abundance changes in both the real-training and synthetic datasets when comparing patients with MAKE to the NE group (e.g., absence of event). A 51.8% overlap was observed in the peptides significantly associated with kidney outcome between the two datasets (**Fig. 5a; Suppl. Table S3**). For the vast majority (97.6%) of these 745 common peptides, the regulation trend was consistent in the real and synthetic datasets, as graphically presented in the heat map analysis (**Fig. 5b; Suppl. Table S3**). The well-established classifier to predict CKD (CKD273)^24–27^ was employed to score both the real-patient and synthetic datasets. A highly significant inverse correlation of the CKD273 score with eGFR in both the real patient (Spearman rho_real-training_ = - 0.394; p = 5.71e-127; **Fig.5c**) and the synthetic datasets (Spearman rho_synthetic_ = - 0.396; p = 4.64e-67; **Fig.5d**), was observed, further confirming the functionality of the algorithm for the synthetic data generation. Similar to the HF2 scores, higher CKD273 scores were correlated with higher probability for developing a future MAKE in both the real- patient (HR = 3.26 ± 0.091; 95% CI; p = 3.65e-38; **Fig. 5e; Suppl. Table S3)** and the synthetic datasets (HR = 5.647 ± 0.155; 95% CI; p = 8.86e-29; **Fig. 5f; Suppl. Table S3**).

**Fig. 5:**
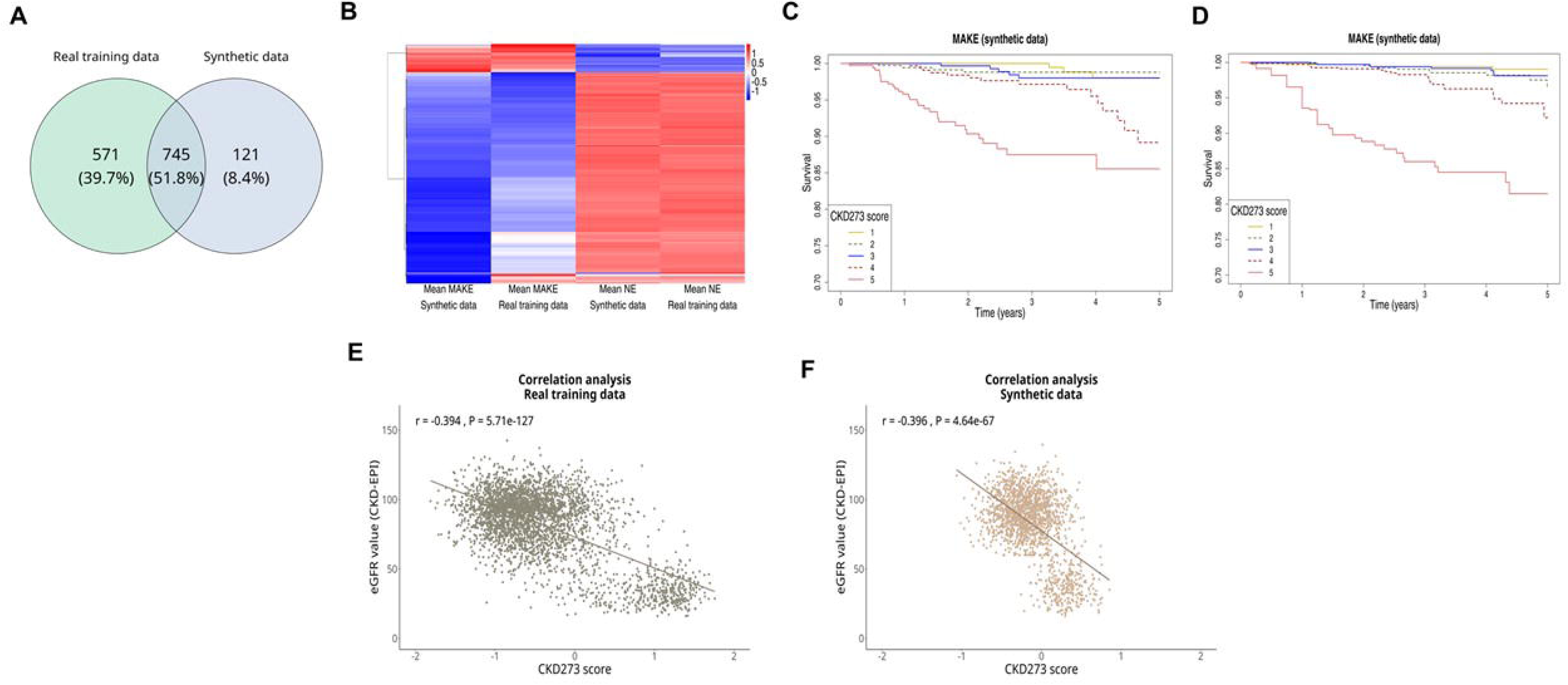
Synthetic data predict chronic kidney disease outcome. **A)** Case control comparison between patients with a kidney event (MAKE) and those that did not develop any disease (Non-event group; NE) **B**) Heat map plot of the most significantly different peptides separating the MAKE and NE groups, in the synthetic and real-patient data. Correlation analysis of CKD273 scores with eGFR in **C)** the real-patient (training) and the **D)** the synthetic datasets. Kaplan-Meier curves for predicting chronic kidney disease event; classifier scores from lowest (1) to highest (5) quintile, for risk of MAKE as assessed by CKD273 real classifier in **E**) the real-patient (training) and **F**) the synthetic datasets.

#### External validation in independent real-patient multicentric datasets

Given the availability of additional external multi-centric cohorts of patients with established chronic kidney disease (CKD, defined as eGFR < 60 mL/min/1.73m²) and controls, we further examined the predictive associations within the synthetic matrix in the context of CKD. The external real-patient validation datasets included CKD of different aetiologies, from three independent cohorts (PersTIgaN, DC-Ren and UPTAKE), as well as subjects with normal kidney function (eGFR > 90 mL/min/1.73m²), in a total of 2,964 datasets (**Table 2**). Independent validation at the single peptide level revealed a reproducible and highly significant association of the rho-values of peptide abundance correlations with eGFR between the external real-validation and synthetic datasets (rho = 0.569; p = 18e-218; **Fig**. **6a**). Stratified data per cohort confirmed the associations in each independent clinical cohort (rho values ranging from 0.466 to 0.516; p < e-137; **Fig. 6c; Supp. Table S4)**. At the multivariate level using the CKD273 classifier^24–27^, highly significant inverse correlation of the CKD273 scores with eGFR was evident, similar to the correlation in the real-patient training data (Spearman rho_synthetic_ = - 0.396; p = 4.64e-67 vs Spearman rho_external_ = - 0.631; p < 2.2e-308; **Fig. 6b**).

**Fig. 6:**
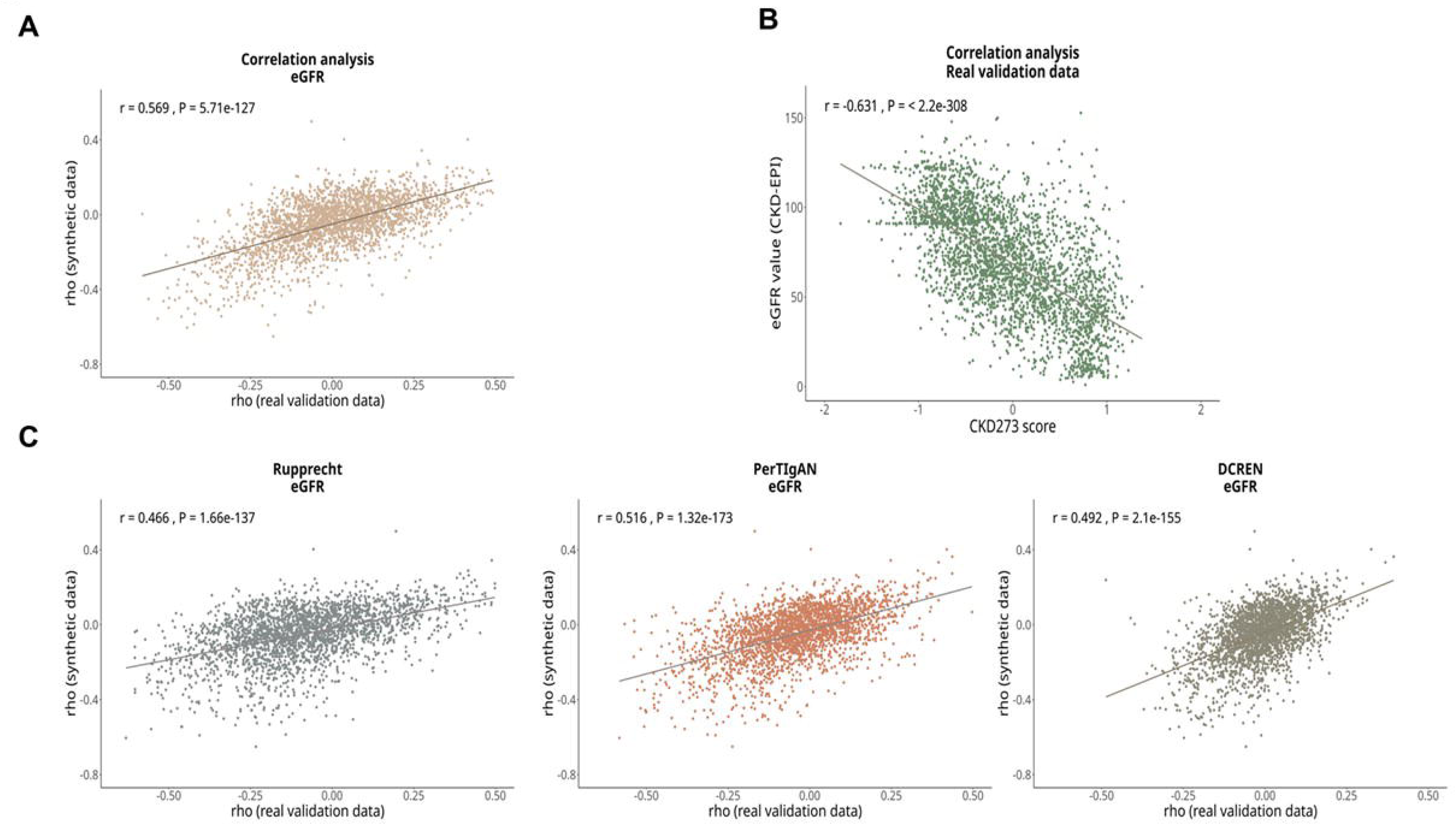
**A)** Correlation plots of spearman rho values between the synthetic and independent real patient data with eGFR**. B)** Correlation analysis of CKD273 scores with eGFR in the synthetic datasets and the second validation real patient dataset; **C)** Stratified analysis per study cohort for the independent real patient data.

**Table 2.** Baseline characteristics of study participants consisting the external validation.

| Characteristic | N = 2,964 external validation patient data - CKD | N = 122 external validation patient data - HF |
| --- | --- | --- |
| **Clinic characteristics** |  |  |
| Age | 62.0 [51.0, 70.0] | 56.5 [44.2, 71.8] |
| Male | 1,734 (58.5) | 48 (54.5) |
| sBP (mm Hg) | 133.0 [123.0, 145.0] | 120.8 [102.2, 142.0] |
| dBP (mm Hg) | 80.0 [70.0, 83.0] | 80.0 [72.8, 86.2] |
| Hypertension | 1,723 (83.6) | NA |
| Diabetes | 1,708 (57.6) | NA |
| eGFR (mL/min/1.73m<sup>2</sup>) | 70.0 [48.2, 93.0] | 73.3 [64.7, 84.7] |
|  |  | HF: 54 (44.3) |
|  |  | Non- HF: 68 (55.7) |
| **CKD** (eGFR < 60 mL/min/1.73m<sup>2</sup>) | 1,096 (54.8) |  |
| **No CKD** (eGFR > 90 mL/min/1.73m<sup>2</sup>) | 904 (30.5) |  |
| Median [IQR]; n (%) |  |  |
| Abbreviations: eGFR, estimated glomerular filtration rate (mL/min per 1.73 m<sup>2</sup>); MI, body mass index; sBP, systolic blood pressure; dBP, diastolic blood pressure |  |  |

#### Synthetic machine learning models validated in real-patient cohorts

Envisaging that a true benefit for a potential application of synthetic data is to make use of high resolution multivariable data towards the identification of biomarkers and biomarker panels without the risk of privacy breaches, we further generated a disease-specific classifier solely based on the synthetic data matrices. Guided by the previous studies developing SVM classifiers related to CKD^24–27^ and HF, we compared patients’ datasets with versus without CKD, or with versus without HF events (**Fig. 7a**). The approach was integrated in two independent SVM development pipelines, one for HF and one for CKD. Based on this, for the synthetic HF-SVM model a case-control statistical analysis was performed within the synthetic datasets, by comparing the patients experiencing an HF event (n = 242) to those without HF and with normal kidney function (eGFR > 90 mL/min/1.73m²; n = 755). As a result, out of 1,083 peptides that were significantly different (adjusted p < 0.05), 676 peptides were integrated in an SVM classifier using a statistical cut-off for highly significant peptides (with p < 10^−6^). Independent validation of the synthetic HF-SVM in an external real patient dataset of 54 patients with and 68 without HF resulted in a significant predictive value depicted by an AUC of 0.803 (0.724-0.881; 95% CI; p < 0.001; **Fig. 7b**, **Supp. Table S5**). Similarly, for developing a synthetic CKD-SVM, a cross-sectional approach was followed within the synthetic datasets, considering 323 patients with CKD or future MAKE that were compared to individuals without a future MAKE and with normal kidney function (eGFR > 90 mL/min/1.73m²; n = 755). Out of 761 significant peptides identified (based on adjusted p value < 0.05), 292 were further considered for the SVM modelling (those highly significant, with p value < 10^−10^). External validation of the synthetic CKD-SVM in a set of 2,000 real-patients data (1,096 CKD patients defined as per eGFR < 60 mL/min/1.73m² and 904 with normal kidney function defined as per eGFR > 90 mL/min/1.73m²), resulted in an AUC value of 0.867 (0.851 - 0.882; 95% CI; p < 0.001; **Supp. Table S5**). Both synthetic SVM models enabled differentiating diseased from non-diseased subjects in external real-patient data in a highly significant manner. In particular for HF sensitivity and specificity estimates were comparable to the previously published HF2 classifier^28^ [Sensitivity (Se) ^28^ = 94%, Specificity (Spe)_Real-HF2_^28^ = 66%; Se_Synthetic-HF_ = 94%; Spe_Synthetic-HF_ = 62%]. As for the synthetic CKD-SVM, a direct comparison of the performance metrics with the CKD273^24–27^ was possible (through comparative ROC curves) as independent data (outside the development/ training data for either synthetic or real SVM) were available. Comparative ROC analysis indicating a slight, yet significant decrease in performance of the synthetic vs the real CKD273 (AUC_Synthetic-CKD_ = 0.867; AUC_Real-CKD273_ = 0.889; p < 0.05).

**Fig. 7:**
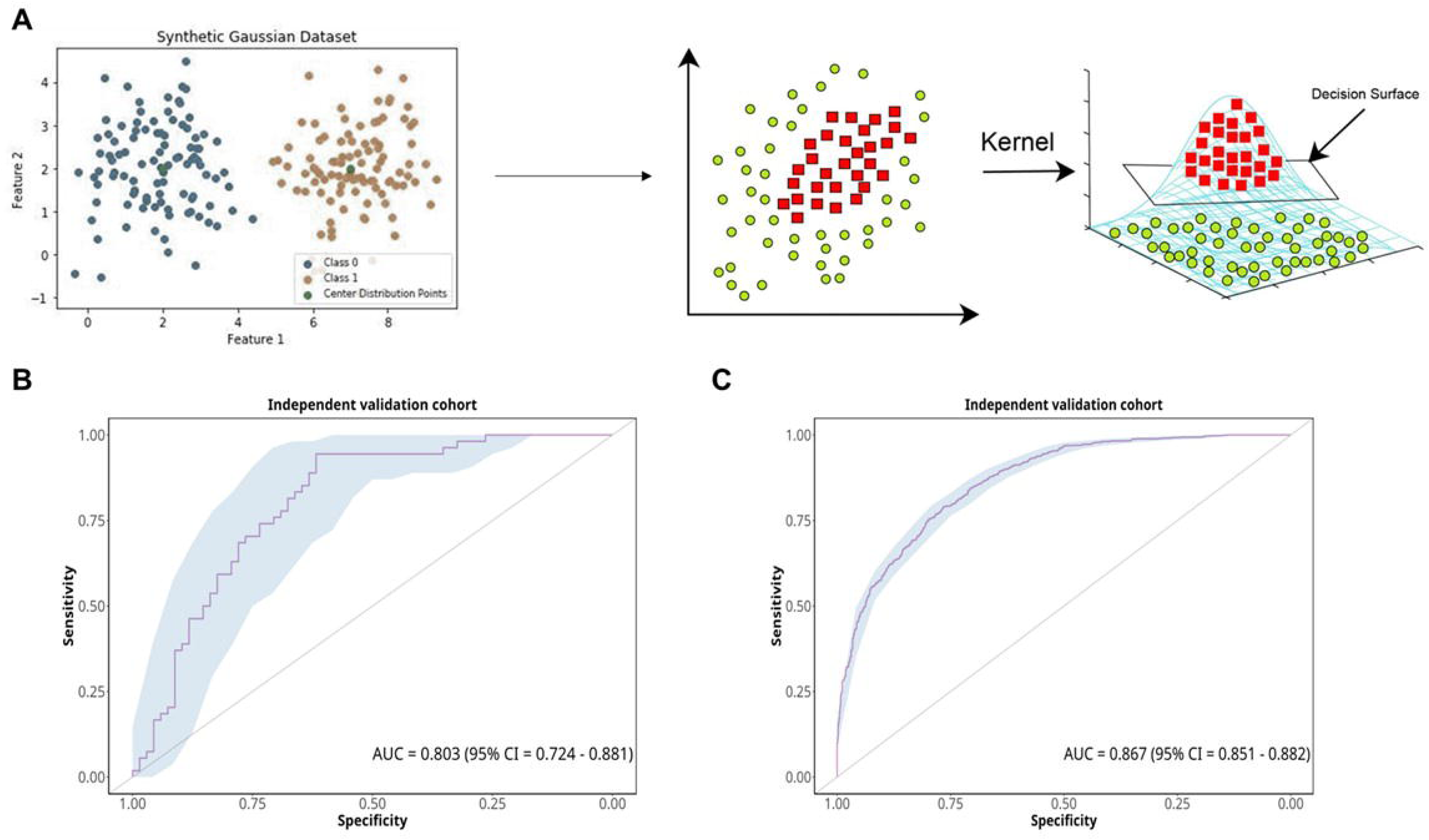
Machine learning models based on synthetic data were reproducibly validated in real-patient cohorts. **A)** Support vector machines were employed for the generation of synthetic classifiers; **B)** A synthetic classifier for detecting HF was validated in external real-patient data; **C)** A synthetic classifier for detecting CKD was also validated in external real-patient data.

## Discussion

Sharing individual health-associated data has become difficult or impossible due to concerns about privacy protection. Data sharing is even more challenging in international projects, when data need to cross borders. This challenge is in part due to diverse regulations in different countries^29^. Restriction in data sharing has undisputedly a substantial negative impact on scientific progress, at the lowest level in delaying progress and requiring additional funds for the respective legal departments involved, but frequently also completely blocking scientific investigations. A potential solution to this problem would be the generation of entirely synthetic patients and data, which per definition cannot be traced back to the original patient(s), and consequently no concerns on privacy protection apply. Application of synthetic data generation models to produce large high resolution molecular synthetic data holds promise to ensure generalisability of models for guiding treatment recommendations across diverse patient populations, and allow for accessing representative datasets without compromising sensitive patient information.

Synthetic data is artificially generated data with the purpose to mimic real data in terms of statistical properties and (predictive) relationships without disclosing any personal information from the original source. In this study, we successfully generated synthetic data that accurately recapitulate both clinical and multi-variable high dimensional abundance data of more than 21,500 variables from a large representative dataset of more than 3,800 patients from ten clinical cohorts investigating kidney disease and/ or heart failure^17, 24, 25, 28, 30–42^. Additional real patient datasets of close to 3,000 subjects from independent clinical studies were used to assess the validity of the generated synthetic data.

Within the real-patient dataset that was used as the basis for the synthetic data generation (training set), a wide range of distributions was developed considering principles of multidimensional Gaussian copulas. By following this approach, the dataset was considered as a visual cloud or multivariate collection of data points, where multiple distributions were assessed and reproduced in the synthetic datasets. In this study, we selected this copula-based method due to its applicability towards high dimensional datasets and its robustness in capturing relationships between variables^19^. Moreover, the method allows for separate modelling of marginal distributions and their correlation structure or matrix, reducing the risk of overfitting and enhancing privacy by avoiding the memorization of any specific patient data — an issue that is frequently present when employing deep learning models^43^. In terms of data privacy, by employing this method that is based on the distribution of all features in the real patient dataspace avoiding any one - to - one synthetic-real patient connection (in other words it is not a patient-centric approach), it is impossible to track back a patient of origin- as such a patient of origin does not exist. Consequently, the risk of patient re-identification is completely eliminated.

Several applications of synthetic data generation methods have been reported in the biomedical field^10^, mainly focusing on replication and/or expansion of clinical and patient metadata (including demographics, single biochemical test results, type of therapies and/or lifestyle data)^44^. Another application of synthetic data generation is on recapitulation of imaging data, for which mainly deep learning and in particular, generative adversarial network (GAN) techniques^9^ have been employed to improve on minor classes (of images) in imbalanced datasets. In terms of high-resolution molecular (omics) data^10^, published studies either report on improvements in modelling and predictive ability of networks and pathways^12^, or improvements of visualisation and dimensionality reduction approaches. A recently published report focused on the improvement of missing values and signal to noise adjustment in proteomics data^45^. However, based on the available literature, there is currently no study demonstrating the generation of synthetic data that retain predictive associations within omics, clinical and outcome variables in a high dimensional dataspace. In terms of preserving patients’ privacy, a recent study reports on synthetic data generation from clinical trial real word patient data including > 1,000 patients, but limited to a maximum of 26 variables (mainly including clinical, demographics and biochemical results)^46^.

In this study, for the first time, fully synthetic data was successfully generated based on large patient cohorts of more than 3,800 individuals, with clinical and follow-up data. This innovative approach represents a significant advancement in the field, particularly given the scale and complexity of the data involved. Our study integrates a large data matrix of 21,568 clinical and omics variables while at the same time successfully recapitulating the predictive capability of the datasets related to disease outcome.

The stability of the algorithm was verified by applying multiple iterations within a bootstrapping approach based on random sampling. Additionally, the robustness of the synthetic data was validated by comparing them and testing the models developed in the synthetic data with independent external real-patient datasets. This additional validation step provided further confirmation that the synthetic data closely mirrors the characteristics and predictive capabilities of real-patient data, reinforcing the reliability of the approach and generated datasets for subsequent analyses. Variability between the results within synthetic and real-patient data is observed; nevertheless, this is in the range of expected variance between the molecular proteomics data from two individuals.

While for the synthetic HF-SVM, performance metrics were similar on the basis of the comparison with previously published validation data^28^, a direct comparison of AUC values was not possible as parts of the real-patient datasets (of 122 patients with or without HF) were previously used for the development of the previously published HF2 classifier. For the synthetic CKD-SVM, comparison with the well-established CKD273 was possible. A slight, yet significant decrease in performance of the synthetic CKD-SVM was observed. This observation is realistic, considering the relative variance that is introduced during the synthetic data generation process, as a compromise to receive de-identified data. Particularly considering the peptidomics data, where the number of missing values is relatively high (as a result of biological but also technical factors), and even though proportion of missing values is retained in the synthetic matrices, variance within the synthetic data generation cannot be avoided.

Despite these minor limitations, considering the presented results, this study sets a new benchmark in the generation and application of synthetic data in clinical research by integrating a vast matrix of variables and validating the synthetic data against multiple real-world datasets. The implications of this work are far-reaching, offering a scalable and privacy-preserving solution for data sharing and subsequent analysis in large cohorts, which could be particularly valuable in personalized medicine and other areas requiring extensive patient data for predictive modelling.

## Methods

### Study participants employed for synthetic data generation

This study included 3,881 datasets from ten clinical cohorts, reported before, i.e. EU-PRIORITY^40^, DIRECT^24^, FLE-MENGHO^47^, CACTI^39^, CardioRen^37^, CAD prediction^34^, Generation Scotland^38^, HOMAGE^31^, SUN-macro^48^, and UZ-Gent^49^. Detailed information on the designs and the methods used in these studies were reported before. The datasets employed in the synthetic data generation pipeline were part of a previously published study^18^. Inclusion criteria were availability of eGFR (calculated using the CKD Epidemiology Collaboration (CKD-EPI) formula), information on MAKE and HF events, and availability of follow-up information^17^. MAKE was defined as > 40% loss in eGFR or kidney failure. An HF event was defined as hospitalization or death from HF. The study was conducted according to the guidelines of the Declaration of Helsinki and all datasets were fully anonymized. The ethics committee of the Hannover Medical School Germany waived ethical approval under the reference number *3116-2016* for all studies involving re-use of data from anonymized urine samples.

### External real-patient datasets for independent validation

For validation, external datasets from patients with CKD (defined as per eGFR < 60 mL/min/1.73m²) and subjects with normal kidney function (eGFR > 90 mL/min/1.73m²), a total of 2,964 datasets were considered (**Table 2)**. These included datasets from diabetic patients enrolled in the Prospective Cohort Study in Patients with T2DM for Validation of Biomarkers (PROVALID) trial^50^. The PROVALID study received ethical approval from the Institutional Review Boards in each participating country, with the Medical University of Innsbruck’s ethics committee providing approval under reference number EK 1188/2020. Additionally, validation data were obtained from the multicentric Personalized treatment in IgA Nephropathy (PersTIgAN) trial^51^, where urine samples and clinical data from biopsy-confirmed IgAN patients were collected from six centers across Europe. This study was also approved by the local Institutional Review Boards at all participating sites. Furthermore, data described by Catanese et al.^52^, involving patients with various CKD etiologies recruited from the Department of Nephrology at Hospital Bayreuth GmbH (Germany), were included. The ethics committee of the Friedrich-Alexander-University Erlangen-Nürnberg approved the nephrological biobank (ethics approval code 264_20 B) and the urinary proteomics analysis (ethics approval code 221_20 B).

### Smaller sized independent cohort for validation of synthetic SVM for HF

Datasets available for 122 subjects that were re-examined for presence of HF due to impaired left ventricular (LV) myocardial function with or without preserved ejection fraction were considered, originally, as part of the Flemish Study on Environment, Genes and Health Outcomes (FLEMENGHO)^28^. This dataset was not considered during the synthetic data generation.

### CE-MS based omics matrix

All CE-MS datasets were from previous studies, all applying the same analytical approach^53–55^. In short, 700 μl of each urine sample were diluted with an equal volume of alkaline buffer containing 2M urea, 10mM NH_4_OH and 0.02% SDS (pH 10.5). Thereafter, the samples were ultrafiltrated using Centrisart ultracentrifugation filters with a cut-off of 20 kDa (Sartorius, Göttingen, Germany) centrifuged at 3000×g. 1,1 ml filtrate was desalted using PD-10 columns (GE Healthcare, Munich, Germany) equilibrated in 0.01% NH_4_OH in HPLC-grade water. The peptide extracts were lyophilised and stored at 4 °C until further use. CE-MS analysis was performed using a P/ACE MDQ capillary electrophoresis system (Beckman Coulter, Fullerton, USA) online coupled to a MicroTOF MS (Bruker Daltonic, Bremen, Germany). Mass spectral ion peaks representing identical molecules at different charge states were deconvoluted into single masses using MosaFinder software^53^. Migration time and ion signal intensity (amplitude) were calibrated and normalised using 29 internal peptide standards with excretion levels generally unaffected by disease^56^. Accuracy, precision, selectivity, sensitivity, reproducibility and stability were reported previously^53, 54^.

### Synthetic data generation pipeline: characteristics of CE-MS datasets

The real-patient dataset that was used for generating the synthetic data comprised of 3,881 patient datasets. The peptide data contained a substantial number of null values, which were defined as missing during the modeling phase. For peptides with a missing signal threshold of less than 40%, Gaussian copula methods were used to synthesize their values, with the model preserving the percentage of missing data in the synthetic dataset. Conversely, peptides with more than 40% missing data were synthesized using mean values derived from the original dataset, as the data were considered insufficient data for Gaussian copula modeling.

### Group specific analysis

To improve analytical accuracy, the Gaussian copula was applied separately to groups based on disease status (kidney disease). This strategy showcases the method’s efficacy in synthesizing complex clinical and peptide data, thereby providing a solid foundation for disease prediction and analysis. This separation was applied to both clinical and peptide data and was necessitated by the discovery that peptides exhibit different distributional characteristics depending on the presence or absence of kidney disease. Based on this analysis the majority of the peptide variables were identified to be right-skewed and follow heavy-tailed distributions, which also showed a substantial impact on the likelihood for kidney disease occurrence (**Supp. Fig. SF1**). By applying the Gaussian copula method separately to each group, data were synthesized to more accurately reflect the unique characteristics of each population. This group-specific approach led to a significant enhancement in the overall quality and reliability of the results.

### Gaussian copula method

For the analysis of peptide and clinical data, an enhanced version of the Gaussian copula, a statistical method that enables joint distribution modelling, was implemented^57^. Briefly, the most suitable marginal distribution was determined through a comprehensive assessment of different distributions (normal, beta, gamma, t, uniform, Gaussian KDE and lognormal) for both continuous and categorical variables. In the case of the categorical variables, the method converts them to numerical based on the proportion within the dataset and assigns them to the interval [0, 1]. Subsequently, the marginal distribution of the values was assigned according to a Gaussian function within the specified interval^57^. Furthermore, three different methods, the Akaike Information Criterion (AIC), the Bayesian Information Criterion (BIC), and the sum of squared errors, were evaluated to ascertain which would be most appropriate for identifying the optimal distribution. Moreover, restrictions were applied during the modelling process to preserve the true nature of the data. Specifically, a constraint was enforced to ensure that diastolic blood pressure values were always lower than systolic blood pressure values. Once the optimal distribution for each variable was determined, the database modelling is defined using the Gaussian copula method, which involves a series of steps:

#### i. Transformation to Uniform Distribution

Each variable is transform into a uniform distribution, defined as follows:

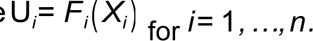

Theorem 1. By the probability integral transform, each *U_i_∼ Uniform*(0, 1).

#### ii. Conversion to Normal Distribution

Once the distribution of the variables is uniform, the inverse cumulative distribution function is applied to convert all variables into a normal distribution:

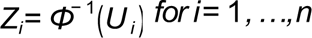

with:

*Φ*^− 1^ is the inverse cumulative distribution function of the standard normal distribution. Theorem 2. Each *Z_i_ ∼ N* (0,1).

#### iii. Estimating the Correlation Matrix

In order to identify the correlation between all transformed variables within the dataset, *Z_i_* ., a correlation matrix *R* was estimated as follows:

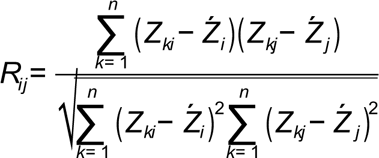

with: *Z_k_i* is the *k* -th observation of the *i* -th transformed variable.

#### iv. Database modelling

A copula is a multivariate probability distribution function C:[0,1]*d* → [0,1] for which the marginal probability distribution of each variable is uniform on the interval [0,1]. According to Sklar’s theorem, any multivariate joint distribution can be written in terms of univariate marginal distribution functions and a copula which describes the dependence structure between the variables using the calculated correlation matrix.

For a *d* -dimensional random vector *X*= (*X*_1_,…, *X*_d_) with marginal cumulative distribution functions *F*_1_,…, *F_d_*, there exists a copula *C* such that for all *x*= (*x*_1_,…, *x_d_*) in *R^d^* :

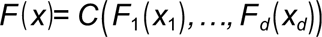

The Gaussian copula is defined as follow,

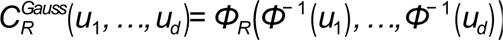

with:

*Φ_R_* is the joint cumulative distribution function of a multivariate normal distribution with mean vector zero and covariance matrix equal to the correlation matrix *R*

*Φ^−^* ^1^ is the inverse cumulative distribution function of a standard normal distribution

#### iv. Generating Samples

Once the model has been defined, the data is synthesized by sampling numerical values from the calculated multi-variable distributions and correlations. These values are then post-processed to be mapped back to the original space, resulting in a representation that mimics the original data, as follows:

1. Generate a sample y from a multivariate normal distribution with mean vector 0 and covariance matrix *R*
2. Transform each component back to the uniform domain: *u_i_*=*Φ(y_i_)*
3. Transform to the original domain using the inverse of the marginal CDFs: *xi* = *F_i_*^−1^(*u_i_*)

#### iv. Bootstrapping

To further ensure stability of the algorithm in generating synthetic datasets and to assess variability caused by, among others, statistical and functional approximation errors, a bootstrapping approach was adopted to allow for estimating sampling distributions using random sampling^22^. As a first step 10,000 patient datasets were generated, including a proportionally same number of HF event, MAKE and NE. For instance, within the 10,000 patient data matrix, 1,253 patients were linked with a future HF event based on the event proportions shown in **Table 1**. Kolmogorov–Smirnov (KS) testing was further applied to compare the peptide distributions, with only 33% of the peptides failing the KS test (significance level 0.05), in majority those with high number of missing peptide abundance values. Consequently, a signal threshold was set to include only peptides with at least 40% non-zero data. The bootstrapping process was repeated for 500 iterations. Additionally, Kullback–Leibler (KL) divergence was applied as a measure for similarity (or difference) between the data distributions^23^ within the real-patient and synthetic datasets. For additional information, please see https://github.com/Atomic-Intelligence/Peptide-synthesis.git.

### Validation of the method

After generating the synthetic dataset, the data distribution for both the synthetic and original datasets was analysed to assess the method’s performance using the “sns.histplot” and “sns.scatterplot” functions. Correlation analysis was performed with the “compare_correlations” package in Python, using default parameters. Principal component analysis (PCA) for the peptidomics matrix and clinical variables was conducted using the “prcomp” function from the “stats” R package under default settings, while UMAP for the peptidomics matrix was performed with the “umap” function from the “umap” R package.

#### Correlation analysis

The correlation analysis of the individual peptides with eGFR and between the CKD273 classifier score and eGFR values was conducted using Spearman’s rank correlation coefficient using the function “cor.test” of the “stats” R package. Correction for multiple testing was based on the Benjamini-Hochberg method using ‘p.adjust’ function of the “stats” R package, and the threshold of significance was an adjusted p < 0.05.

### Statistical analysis

All statistical tests were performed using the R statistical software (R version 4.1.0, R Foundation for Statistical Computing, Vienna, Austria)^56^. The statistical comparison between cases and controls was conducted using a peptide frequency threshold of 40% in at least one group. P-values were calculated using a Wilcoxon test with the “wilcoxon.test” function and multiple testing correction was applied using the Benjamini-Hochberg method using the ‘p.adjust’ function from the “stats” package. Significant peptides, determined by an adjusted p < 0.05, were compared between the real and synthetic datasets using the “ggvenn” function and the heatmaps were generated using the “pheatmap” package.

### Peptide-based classifiers and prediction of events

The predictive capacity of the classifiers CKD273^23–26^ and HF2^27^ was assessed for predicting future events. The scores for each classifier were calculated using a SVM algorithm, integrated into the MosaDiagnostics software^54^, area under the receiver operating characteristic (ROC) curve (AUC) of the generated models were performed using the “roc” function from the “pROC” package. The Kaplan-Meier estimator was applied to assess the association of longitudinal survival using the HF2^56^ and CKD273^23–26^ classifier scores. Corresponding HR were estimated using Cox regression models and log-rank tests were used to assess the hypothesis of no group differences in hazard functions. Survival analyses were carried out using the package “survival” and all the graphs were done with the package ‘ggplot2’.

## Data availability

The data that support the findings of this study are available from the corresponding author upon reasonable request.

## Code availability

The synthetic data generation package was written in Python. The script is freely available under MIT license (https://github.com/Atomic-Intelligence/Peptide-synthesis.git).

## Ethics declarations

HM is the cofounder and co-owner of Mosaiques Diagnostics (Hannover, Germany). MAJC, MF, AL and JS are employees of Mosaiques Diagnostics. TK is the cofounder and co-owner Atomic Intelligence (Zagreb, Croatia). SK, EA, VD, and DV are employees of Atomic Intelligence. All other authors declare no competing interests.

## Acknowledgments – Funding information

The Authors sincerely thank Mr Igor Golovko for his invaluable support with data transformation. MAJC holds a doctoral grant through the DisCo-I project that has received funding from the European Union’s Horizon Europe Marie Skłodowska-Curie Actions Doctoral Networks - Industrial Doctorates Programme (HORIZON – MSCA – 2021 – DN-ID) under grant agreement No 101072828, funded by the European Union. This study was in part funded by UPTAKE (urinary proteome analysis for the prediction of type, extent, prognosis, and therapeutic response of acute and chronic kidney diseases), funded by the Bundesministerium fur Bildung und Forschung (BMBF; Federal Ministry of Education and Research) in the program “Translational projects in personalized medicine” under the grant numbers 01EK2105A, 01EK2105B, and 01EK2105C, by SIGNAL funded by BMBF (grant number 01KU2307) and by Austrian Science Fund (FWF, Project number I 6464, Grant-DOI 10.55776/I6464), by Accurate-CVD (ZIM-KK5560002AP3) funded by the by the BMWK (Federal Ministry for Economic Affairs and Climate Protection), ProSTRAT-AI (01DS23014) funded by the BMBF (Federal Ministry of Education and Research), PERMEDIK COST Action, supported by COST (European Cooperation in Science and Technology) grant no.CA21165 and DC-ren (Horizon 2020 research and innovation programme under grant agreement No 848 011) and MULTIR (HORIZON-MISS-2023-CANCER-01-01; project number: 101136926) both funded by the European Commission. Views and opinions expressed are however those of the author(s) only and do not necessarily reflect those of the European Union. Neither the European Union nor the granting authority (HORIZON – MSCA – 2021 – DN-ID, Horizon 2020 research and innovation programme and HORIZON-MISS-2023-CANCER-01-01) can be held responsible for them.

