## Supplementary figures and images for "A synthetic data generation pipeline to reproducibly mirror high-resolution multi-variable peptidomics and real-patient clinical data"

### Supp. Fig. SF1

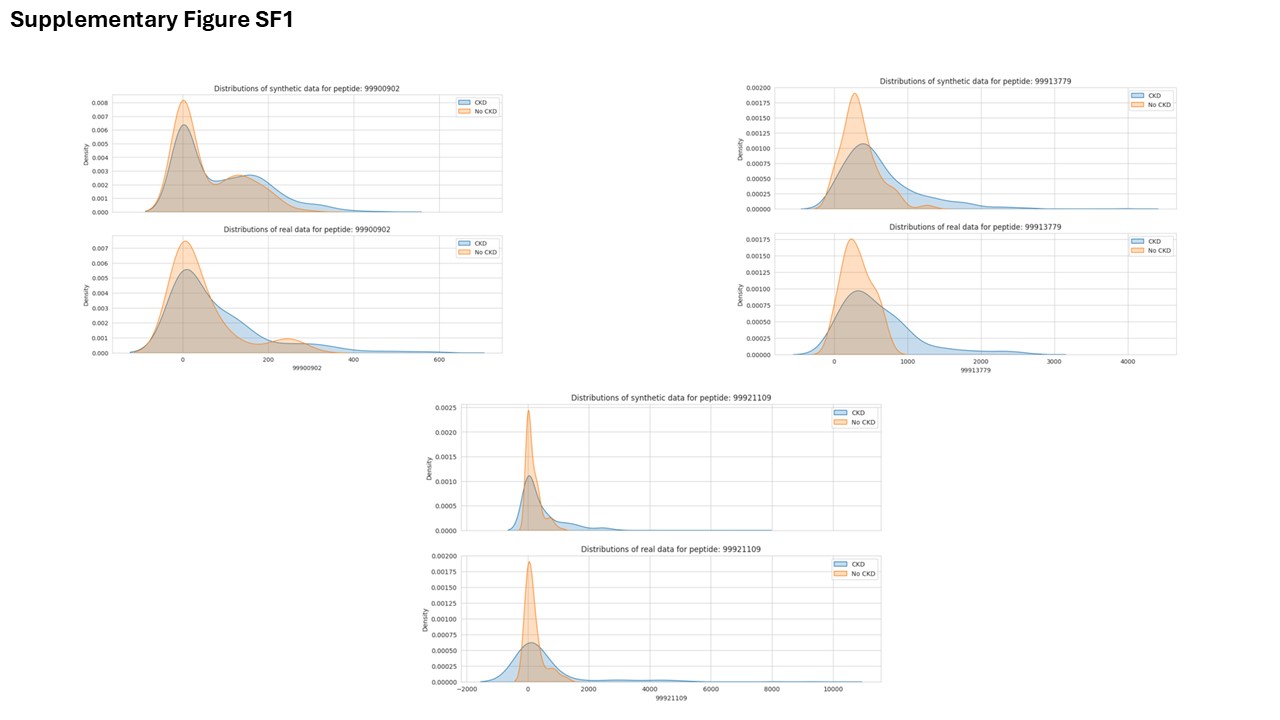

### Supp. Fig. SF2

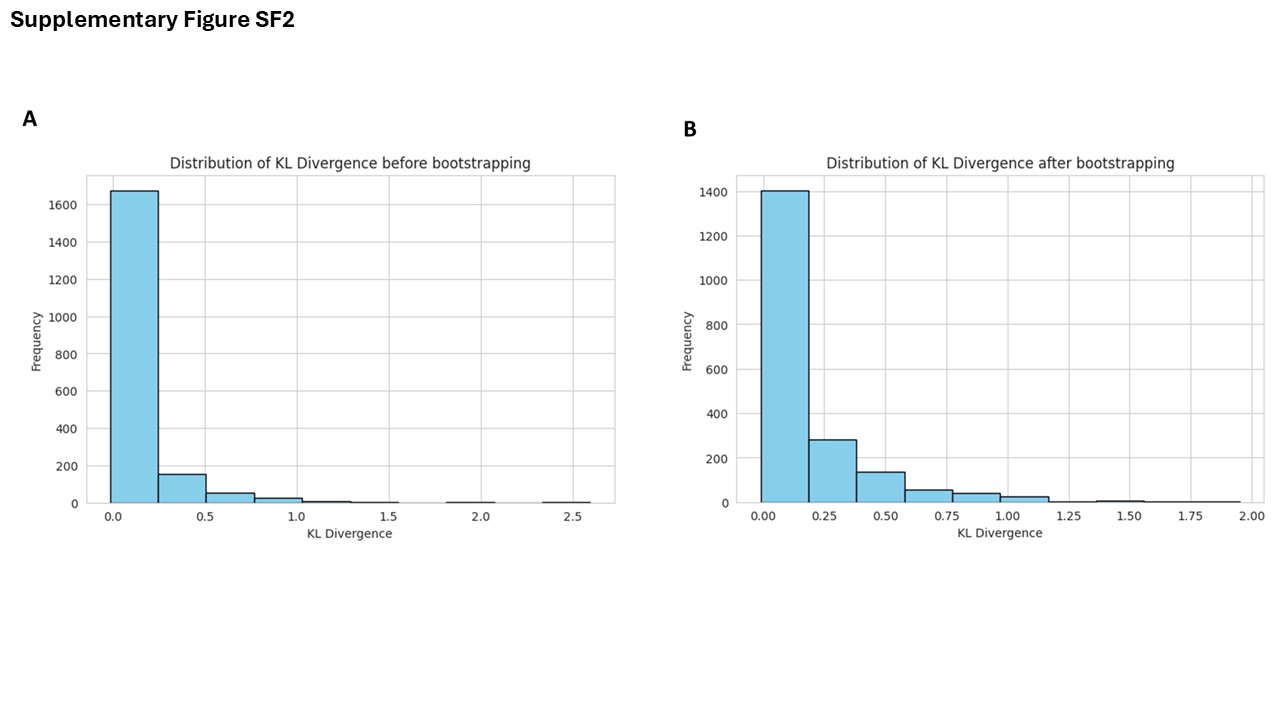

### Supp. Fig. SF2

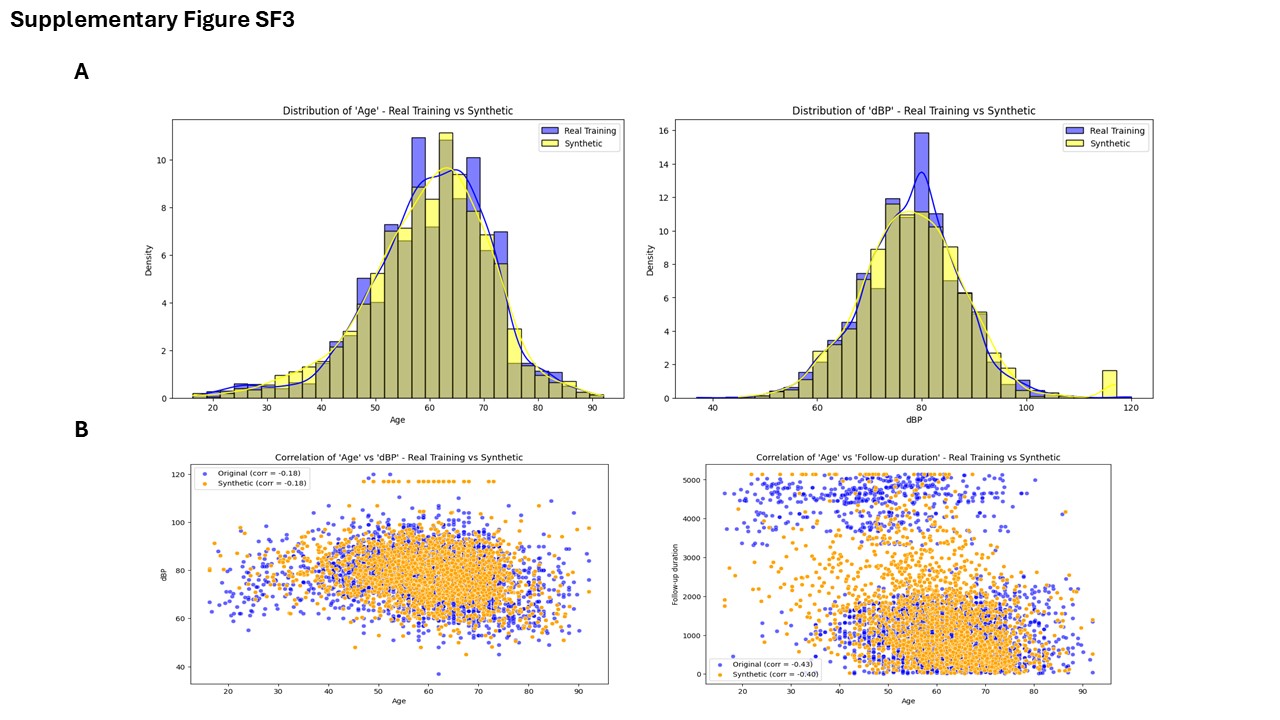
